# Living Environments and Mental Health: the environMAP database

**DOI:** 10.64898/2026.05.15.26353275

**Authors:** Paul Renner, Elli Polemiti, Marcel Jentsch, Jamie R. Banks, Dennis Cleff, Sebastian Siehl, Marco Dallavalle, Tristram Lett, Christoph Buck, Stefanie Castell, Jonas Frost, Hans Grabe, Thomas Keil, Volker Harth, Robyn Kettlitz, Lilian Krist, Michael Leitzmann, Rafael Mikolajczyk, Nour Naaouf, Nadia Obi, Annette Peters, Alexandra Schneider, Kathrin Wolf, Frauke Nees, Sven Olaf Twardziok, André Marquand, Sören Hese, Kerstin Schepanski, Gunter Schumann, the environMENTAL consortium

**Author notes:** Equally contributing authors (Paul Renner, Elli Polemiti).

## Abstract

Environmental exposures are increasingly examined in relation to mental health, yet large-scale epidemiological analyses remain constrained by fragmented geospatial data, heterogeneous spatial and temporal resolutions, and privacy-preserving linkage requirements, limiting systematic investigation of multiple environmental domains at the population level.

We present environMAP, a harmonised set of analysis-ready environmental exposure layers derived from open, global sources. environMAP spans the built environment, green and blue spaces, light exposure (solar radiation and night-time light), terrain, weather and extremes, and air pollution. We document data provenance, spatial buffers, preprocessing, projection alignment, and metadata, and provide a reproducible workflow for privacy-preserving linkage to cohort residential locations. To demonstrate utility, we linked environMAP to >200,000 adults in the German National Cohort (NAKO) and summarised self-reported lifetime doctor-diagnosed depression across exposure gradients using sex-stratified descriptive analyses. Gradients were interpretable and broadly consistent with prior evidence, supporting feasibility, scalability, and hypothesis generation. The framework is adaptable to other outcomes, cohorts, and regions.

## Introduction

Environmental exposures—the air we breathe, the light we experience, and the physical structure of our surroundings—shape both physical and mental health. Air pollution, temperature extremes, noise, urban form, and access to green and blue spaces have each been linked to depression, anxiety, and other psychiatric outcomes, suggesting that environmental stressors contribute meaningfully to population mental well-being^1^. At the same time, how multiple environmental domains combine and interact to influence mental health remains incompletely understood^2^.

A growing epidemiological literature reports associations between air pollutants and depression or anxiety^3,4^, as well as between access to green and blue spaces and lower psychological distress^5,6^. Urbanicity, heat exposure, and artificial light at night have similarly been implicated in mood and stress-related disorders^7,8^. Taken together, these findings indicate that environmental factors matter for mental health, but they also reveal substantial variation in exposure definitions, spatial scales, and analytical approaches across studies.

Methodological challenges have limited synthesis and comparability. Environmental indicators differ widely in spatial and temporal resolution, coordinate systems, units, and update frequency, and are released by multiple agencies in heterogeneous formats. Integrating multiple environmental domains into a single, reproducible analysis therefore remains technically demanding for many research groups. As a result, many studies focus on single exposures or narrow domains, limiting insight into how environmental stressors cluster, co-vary, or jointly shape mental-health outcomes^2^.

Practical constraints further restrict analytical scalability. Linking detailed environmental data to individual residential locations raises privacy concerns, particularly in large population cohorts. Few studies have established transparent and reproducible workflows that enable multidomain environmental linkage while safeguarding participant confidentiality. At the same time, environmental stressors typically exert modest effects at the individual level^9^, underscoring the need for large, well-characterised cohorts and harmonised exposure assessment.

Recent advances in open-access geospatial products derived from satellite observations and global reanalyses have transformed the environmental data landscape. High-resolution information on air quality, weather, vegetation, hydrology, and urban form is now available across large geographic areas and extended time periods. These developments create unprecedented opportunities for environmental mental-health research, but only if the underlying data can be systematically integrated and linked to health studies in a reproducible manner.

To address these challenges, we developed environMAP, (Mapping Application Process for environmental datasets) (https://prenner-eo.github.io/environMAP/), a curated and harmonised framework within the environMENTAL project (https://www.environmental-project.org/) that enables scalable, privacy-preserving linkage of multidomain environmental data to health cohorts. environMAP organises existing geospatial datasets in a unified structure, documents data provenance, standardises spatial processing, and ensures traceability to public sources. Linkage can be performed centrally within a secure high-performance computing (HPC) environment or locally by participating cohorts, providing flexibility while maintaining data protection.

In this paper, we introduce environMAP and demonstrate its application using data from the German National Cohort (NAKO)^10,11^. The analyses presented here are illustrative and intended to demonstrate the utility of environMAP for large-scale, multidomain environmental mental-health research. We describe the environmental domains included, the procedures used for data preparation and linkage, and an example analysis of lifetime depression across selected environmental gradients. Although the demonstration focuses on mental health, the framework is applicable to a wide range of health outcomes and research contexts.

## Results

### environMAP: harmonised multidomain environmental data

environMAP integrates a comprehensive set of open-access geospatial datasets that characterise environmental dimensions relevant to mental-health research. The framework currently includes more than 130 environmental variables derived from ten major datasets spanning build structures ^12^, vegetation^13,14^, hydrology ^15^, night-time lights^16^, solar radiation^17^ (and a novel solar radiation dataset provided by Julien Radoux), topography^18^, weather and weather extremes ^19^, and air quality^20^ (Supplement A).

Datasets were selected based on quality-controlled preprocessing, spatial coverage, and long-term availability, enabling harmonised analyses across cohorts and regions. Data were retrieved directly from data providers or via official distribution platforms such as the Copernicus browser^21^. All layers were standardised using consistent projections, spatial resolutions, and file structures, and prepared for analysis within a structured server-based storage system.

This infrastructure supports secure, high-capacity hosting for diverse geospatial layers and enables controlled access, versioning, and efficient data sharing among consortium partners or external researchers. Machine-readable metadata and open formats (e.g. GeoTIFF, NetCDF) ensure interoperability with geographic information systems, statistical software, and modelling pipelines. These design choices facilitate transparent reuse and reproducibility across studies.

environMAP is currently implemented in divers cohort studies, including NAKO^10,11^ (Germany), KiGGS^22^ (Germany), MoBa^23^ (Norway), UK Biobank^24^ (United Kingdom), and IMAGEN^25^ (France, Ireland, Germany, United Kingdom), the Chinese CHIMGEN^26^, ZIB (https://zib.fudan.edu.cn/index) and Yufu ^27^ cohorts as well as the Indian cVEDA^28^. Together, these cohorts comprise hundreds of thousands of participants across adult population samples, birth cohorts, and mental health–focused studies, illustrating the framework’s applicability across diverse study designs and populations.

### Privacy-preserving linkage of environmental exposures to cohort data

A central feature of environMAP is the ability to link environmental exposures to individual cohort participants while safeguarding privacy. Two complementary linkage workflows were established to accommodate different cohort infrastructures, governance models, and regulatory requirements.

In the central linkage workflow, an authorised researcher from the environMENTAL consortium performs linkage within the secure HPC environment. Cohort data managers provide pseudonymised identifiers and participant coordinates, optionally rounded to 100 m to enhance privacy protection. Environmental exposures are assigned using standardised scripts, and only de-identified exposure tables are returned to the cohort. All files containing location data are subsequently removed.

In the local linkage workflow, authorised cohort personnel access the HPC environment directly, where a dedicated project folder contains all required geospatial layers, variable dictionaries, and scripts. The linkage is executed locally with technical support from the environMENTAL data management team. The Environmental Data Unit of NAKO exemplifies this workflow, linking environMAP layers to the cohort’s 100 × 100 m residential grid without transferring identifiable location data outside the cohort infrastructure.

Both workflows yield identical exposure definitions and fully reproducible results. This flexibility allows cohorts to select a linkage approach aligned with their governance structures while maintaining harmonisation across studies.

### Spatial distribution of environmental exposures across Germany

To contextualise the demonstration analysis, figure 1 presents national maps of selected environMAP indicators across Germany. Building volume delineates dense urban structures concentrated in major metropolitan regions, contrasting with extensive low-density rural landscapes. Tree cover outlines forested regions and major urban green spaces, while blue-space proximity reflects the sparse distribution of large surface waters, with higher values near lakes, reservoirs, and major river systems.

**Fig. 1:**
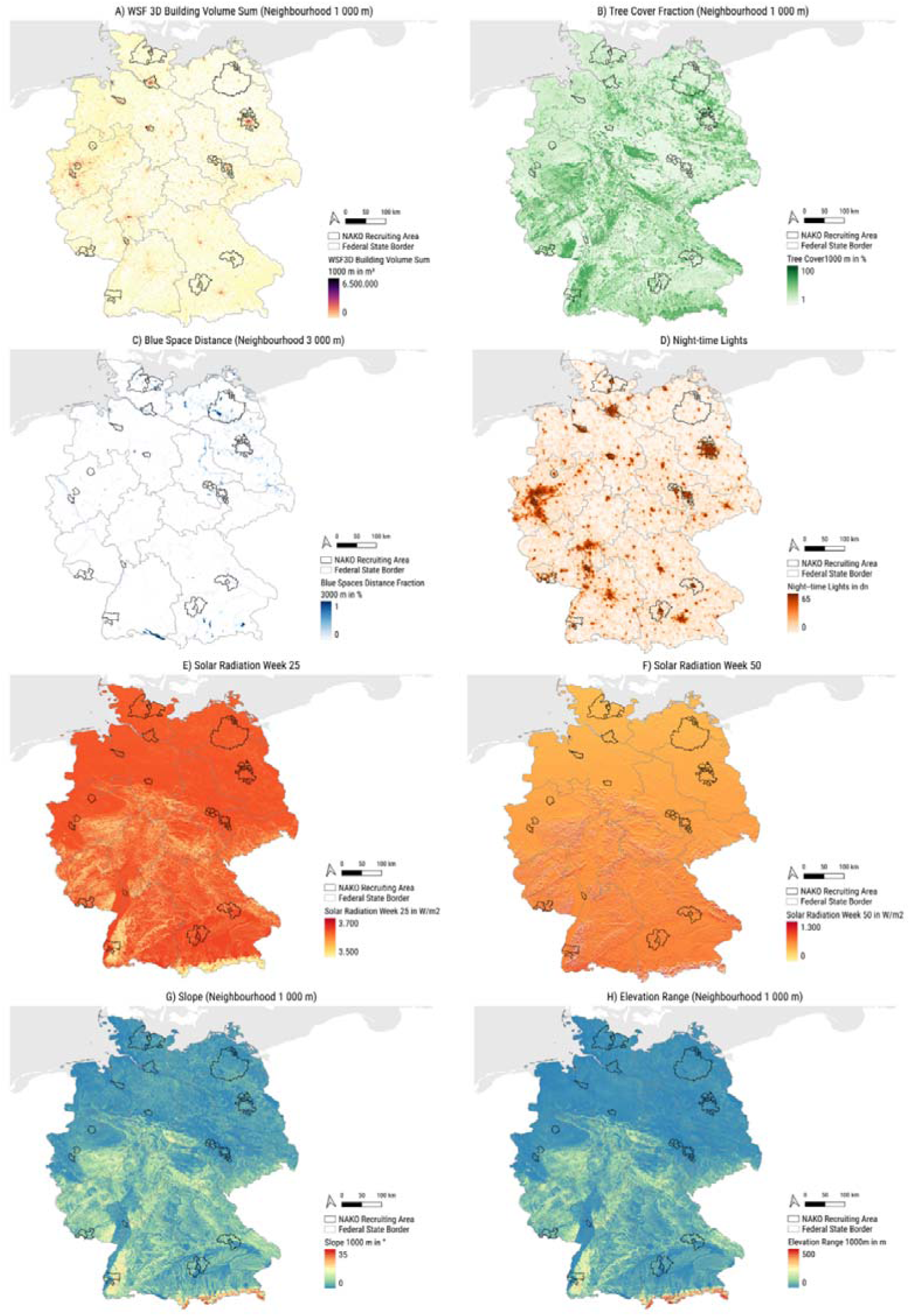
Spatial distribution of environmental indicators across Germany. Panels show national maps of neighbourhood-scale environmental exposures derived for NAKO participants: (A) building volume (1 km diameter), (B) tree-cover fraction (1 km diameter), (C) blue-space proximity (3km diameter), (D) night-time light intensity (1 km diameter), (E) week 25 solar radiation (300 m), (F) week 50 solar radiation (300 m), (G) terrain slope (1 km), and (H) elevation range (1 km diameter).

Night-time light intensity traces highly developed and populated corridors, mirroring national patterns of infrastructure and settlement. Solar radiation shows relatively uniform values in summer but greater heterogeneity in winter across NAKO recruitment regions. Topographic indicators, including terrain slope and elevation range, capture Germany’s physical variability, distinguishing flat northern plains from more rugged southern terrain.

Together, these maps illustrate substantial spatial heterogeneity and partial correlation across environmental domains. Some indicators, such as building volume and night-time light, show strong spatial alignment, whereas others, vary independently. This reinforces the importance of multidomain exposure assessment rather than reliance on single indicators as proxies for environmental context.

### Distribution of environmental exposures by depression status

We linked environMAP exposures to 203,121 NAKO participants with valid address information and complete depression status. Lifetime depression was defined by self-report of a doctor- or psychotherapist-diagnosed depressive disorder.

Table 1 summarises neighbourhood-scale environmental indicators by depression status and sex on individual values. Women reported higher lifetime depression prevalence than men, consistent with established population patterns^29^. Participants with lifetime depression reported higher median exposure to built-environment density, as measured by WSF3D building volume, and higher night-time light intensity compared to those without depression. These patterns were consistent across sexes and suggest a modest tendency for participants with depression to reside in more urbanised neighbourhoods. In contrast, green space (tree cover), blue space proximity, solar energy, slope and elevation showed negligible differences between depression groups, with largely overlapping interquartile ranges across all strata.

**Table 1:** Summary of environmental indicators by lifetime depression status and sex in the NAKO cohort.

| Environmental Indicators | Total Min | Total Max | Total Population Median (IQR) |  | No Depression Median (IQR) |  | Lifetime Depression Median (IQR) |  |
| --- | --- | --- | --- | --- | --- | --- | --- | --- |
|  |  |  | Men<br>n=100,758 | Women<br>n=102,363 | Men<br>n=90,115 | Women<br>n=83,253 | Men<br>n=10,643 | Women<br>n=19,110 |
| WSF3D Building Volume, m<sup>2</sup> | 0.00 | 10,331,845 | 495340.00<br>(927374.75) | 496207.00<br>(919632.00) | 484084.00<br>(906751.50) | 483047.00<br>(899665.50) | 598135.00<br>(1052745.50) | 560560.00<br>(1007385.50) |
| Tree cover, % | 5.27 | 65.68 | 16.24<br>(9.48) | 16.44<br>(9.46) | 16.18<br>(9.51) | 16.39<br>(9.53) | 16.63<br>(9.16) | 16.69<br>(9.19) |
| Blue Spaces | 0.00 | 0.60 | 0.00<br>(0.01) | 0.00<br>(0.01) | 0.00<br>(0.01) | 0.00<br>(0.01) | 0.00<br>(0.01) | 0.00<br>(0.01) |
| Night-time Lights | 0.00 | 63.00 | 55.00<br>(31.00) | 55.00<br>(31.00) | 54.00<br>(32.00) | 54.00<br>(32.00) | 57.00<br>(25.00) | 56.00<br>(27.00) |
| Sun Energy (Week 25), W/m<sup>2</sup> | 2,127.00 | 3,666.00 | 3637.00<br>(9.00) | 3637.00<br>(9.00) | 3637.00<br>(9.00) | 3637.00<br>(9.00) | 3637.00<br>(9.00) | 3637.00<br>(9.00) |
| Sun Energy (Week 50), W/m<sup>2</sup> | 0.00 | 1793.00 | 537.00<br>(178.00) | 536.00<br>(175.00) | 537.00<br>(177.00) | 536.00<br>(176.00) | 538.00<br>(178.00) | 535.00<br>(173.00) |
| Slope, ° | 0.50 | 27.18 | 3.25<br>(1.82) | 3.26<br>(1.81) | 3.23<br>(1.83) | 3.24<br>(1.81) | 3.37<br>(1.78) | 3.34<br>(1.77) |
| Elevation Range, m | 3.82 | 409.98 | 22.46<br>(14.42) | 22.42<br>(14.45) | 22.45<br>(14.48) | 22.45<br>(14.61) | 22.52<br>(14.00) | 22.28<br>(13.69) |

IQR, interquartile range. Environmental indicators were defined at 1,000m neighbourhood scale, except blue-space proximity (3,000m).

Together, these distributions illustrate the heterogeneous residential contexts of NAKO participants and demonstrate that environMAP-derived exposures are available and interpretable across the full cohort, providing a suitable basis for further multivariable analyses.

### Exploratory exposure–depression gradients across environmental domains

To visualise continuous exposure-outcome relationships, participants were ranked by each environmental variable and grouped into sex-specific bins of 1,000 individuals. Within each bin, the percentage of participants with lifetime depression was calculated, and the mean exposure value was assigned for plotting. This binning strategy stabilises prevalence estimates, smooths random variation, and facilitates comparison across rural-urban exposure gradients.

Exploratory linear and quadratic regressions were fitted to the grouped data to provide a descriptive summary (Supplement B). As this approach operates on grouped summaries rather than individual-level observations, the resulting regression fits are intended as descriptive visual aids and should not be interpreted as effect estimates. Quadratic terms were included to capture nonlinear patterns that may arise across environmental gradients.

Depression prevalence increased from low building volume to moderate densities and then plateaued at higher values, suggesting a nonlinear pattern across the urban gradient and aligning with prior evidence on higher depression risk in medium-density settings ^30,31^. Night-time light intensity showed one of the strongest gradients, with higher illumination associated with higher depression prevalence in both sexes.

Associations with tree cover and blue-space proximity were comparatively weak and nonlinear. Importantly, this likely reflects limited within-cohort exposure contrast for some indicators rather than evidence of no relationship. For green space, associations between tree cover and depression were weak and non-linear, with slightly higher depression percentages at low-to-intermediate greenness and greater variability at the lowest and highest levels; overall, these patterns are consistent with evidence suggesting modest protective effects of green space on depression ^32^.

For blue space, most NAKO participants had near-zero values even using the larger 3 km neighbourhood definition (Table 1), constraining the ability to detect gradients despite prior reports of modest benefits of green or blue environments ^33–35^. Likewise, solar radiation showed low spatial variability within each season across the NAKO study setting. Summer (week 25) values were almost uniform across Germany, and winter (week 50) values, while more heterogeneous than summer, still varied only modestly across recruitment areas, so both seasonal indicators explained little variation in depression prevalence.

Terrain slope showed a modest nonlinear association, whereas elevation range was not meaningfully related to depression; similar slope-related patterns have been reported where steeper terrain may reduce mobility and attenuate potential mental-health benefits ^36^.

Overall, these exploratory gradients demonstrate that environMAP-derived exposures produce coherent, directionally plausible population-level patterns, particularly for urbanicity-related indicators. While the observed gradients are descriptive and do not support causal inference, they indicate that environMAP exposures are sufficiently sensitive to detect meaningful variation at population scale, supporting their use in multivariable analyses.

### Multidomain environmental gradients capture distinct aspects of residential context

Beyond individual gradients, the grouped analyses indicate that different environmental domains capture partially overlapping but non-redundant aspects of residential context. Urbanicity indicators, such as building volume and night-time light intensity, showed broadly similar gradient shapes but differed in magnitude and curvature, whereas natural-environment indicators were generally flatter and more variable. Together, these descriptive results show that environMAP-derived exposures provide interpretable population-level contrasts across environmental domains and offer a structured basis for hypothesis generation and subsequent multivariable analyses.

### Robustness of environmental gradients across sex and spatial scale

The sex-stratified presentation further illustrates the descriptive utility of environMAP for comparing environmental gradients across subgroups. Although women consistently showed higher percentages of lifetime depression across exposure ranges, the overall shape of the gradients was broadly similar between sexes for most indicators, suggesting that the environmental patterns captured here are not driven solely by sex-specific distributional differences. This visual consistency supports the use of environMAP exposures for comparative analyses and for future studies examining potential effect modification in larger or more heterogeneous samples. In addition, the generally stable gradient patterns across most exposure ranges suggest that the selected neighbourhood definitions provide interpretable contextual measures at the population level. Where gradients appeared weak or largely flat, such as for solar radiation or blue-space proximity, this more likely indicates limited spatial variation in these indicators within the present application than a lack of utility of the framework itself. Taken together, these observations show that environMAP enables structured descriptive comparison of environmental patterns across domains, subgroups, and neighbourhood definitions, and can serve as a coherent basis for subsequent multivariable, longitudinal, and sensitivity analyses.

## Discussion

This study presents environMAP as a harmonised and scalable framework for integrating multidomain environmental data into mental-health research. By consolidating diverse geospatial datasets into an analysis-ready infrastructure, environMAP addresses a key technical barrier that has limited systematic investigation of environmental determinants.

The application to the NAKO cohort demonstrates that environMAP enables reproducible, privacy-preserving linkage of environmental exposures at national scale and supports stable, interpretable summaries of exposure patterns. The observed exposure–depression gradients—such as higher depression prevalence in more urbanised and illuminated environments and weaker or context-dependent associations for greenness and blue space—are consistent with previous studies^31,37–44^ and support the internal coherence of the framework.

A persistent challenge in environmental health research is the mismatch of spatial and temporal resolution across datasets. Atmospheric composition layers are often available at coarse spatial scales, whereas land-cover and built-environment metrics can reach tens of metres. Such mismatches can dilute spatial contrasts and complicate joint interpretation across domains. environMAP provides harmonised exposure definitions but does not eliminate the need for careful scale selection tailored to specific research questions.

Temporal variability poses additional challenges. Climatic and pollution exposures vary seasonally and interannually, and extreme events may exert disproportionate effects on mental health. Incorporating higher temporal resolution products will enable more detailed examination of exposure windows and temporal dynamics. The complementary CLUES framework (https://bih-dmbs.github.io/CLUES/) extends environMAP by automating ingestion of higher-resolution weather, atmospheric, and socioeconomic datasets, further expanding analytical flexibility.

Environmental exposures operate within broader social and behavioural contexts. Integrating environMAP with mobility and time-use data, such as those collected through the StreetMind initiative^45,46^, offers a pathway toward activity-based exposure modelling that better reflects lived experience and reduces misclassification.

Analytically, multidomain environmental research must contend with spatial autocorrelation, correlated exposures, and high dimensionality^47–49^. environMAP facilitates such work by providing consistent exposure definitions across cohorts and spatial scales, enabling sensitivity analyses, multi-exposure modelling, and cross-cohort comparison.

From a mental-health research perspective, environMAP addresses a growing need to move beyond isolated exposure analyses toward more integrative representations of environmental context. Mental-health outcomes such as depression are unlikely to be shaped by single environmental factors in isolation, but rather by constellations of co-occurring exposures related to urban form, natural environments, weather, and infrastructure. By providing harmonised access to these domains within a single framework, environMAP facilitates analyses that can examine how environmental features cluster spatially, how gradients differ across domains, and how combinations of exposures characterise different residential contexts. This capability is particularly relevant for comparative research across cohorts and regions, where inconsistent exposure definitions have historically limited synthesis. The framework therefore supports a shift from fragmented, single-domain analyses toward more holistic descriptions of environmental context in mental-health research.

The scalability of environMAP is also relevant for future applications in prevention-oriented and policy-relevant research. Harmonised environmental indicators allow mental-health outcomes to be examined in relation to modifiable features of the built and natural environment, such as urban density, access to green space, or night-time illumination. Although the present analyses are illustrative, the framework enables consistent exposure definition across time and place, supporting monitoring of environmental change and comparison across jurisdictions. In addition, environMAP provides a common foundation for integrating environmental data with other modalities increasingly used in mental-health research, including socioeconomic indicators, behavioural data, and longitudinal health records. By lowering technical barriers to such integration, environMAP creates opportunities for more comprehensive analyses of how environmental context intersects with social and individual factors across the life course.

In summary, environMAP provides a robust and flexible platform for large-scale environmental mental-health research. By lowering technical barriers and promoting harmonisation, it enables more integrated analyses of environmental context and supports hypothesis-driven, comparative, and longitudinal studies across diverse populations.

## Data Availability

All geospatial data produced in the present study are available upon reasonable request to the authors.

## Acknowledgement

Funded by the European Union (Grant agreement No 101057429). Complementary funding was received by UK Research and Innovation (UKRI) under the UK government’s Horizon Europe funding guarantee (10131373 and 10038599) and the National Key R&D Program of Ministry of Science and Technology of China (MOST 2023YFE0199700). Views and opinions expressed are however those of the author(s) only and do not necessarily reflect those of the European Union, the European Health and Digital Executive Agency (HADEA), UKRI or MOST. Neither the European Union nor HADEA nor UKRI nor MOST can be held responsible for them.

This work supports the Earth Brain Health Commission.

This project was conducted with data (Application No. NAKO-784) from the German National Cohort (NAKO) (http://www.nako.de). The NAKO is funded by the Federal Ministry of Education and Research (BMBF) [project funding reference numbers: 01ER1301A/B/C, 01ER1511D, 01ER1801A/B/C/D and 01ER2301A/B/C], federal states of Germany and the Helmholtz Association, the participating universities and the institutes of the Leibniz Association. We thank all participants who took part in the NAKO study and the staff of this research initiative.

Special thanks to Julien Radoux, for providing the solar radiation dataset.

Produced using Copernicus WorldDEM-30 © DLR e.V. 2010-2014 and © Airbus Defence and Space GmbH 2014-2018 provided under COPERNICUS by the European Union and ESA; all rights reserved.

## Supplement A: Geospatial Data Collection environMAP

1 Supplement A: Geospatial Data Collection environMAP
| Variable Name | Data Source | Spatial Resolution | Neighbourhood (Diameter) [m] | Temporal Coverage | Units |
| --- | --- | --- | --- | --- | --- |
| Grey Spaces: World Settlement Footprint (WSF) 3D |  |  |  |  |  |
| Average Building Height | 12 | 90 m | None | static with data from 2010-2015 | dm |
| Building Fraction |  |  | None |  | % |
| Total Building Area |  |  | None |  | m² |
| Total Building Volume |  |  | 90/1000/2000 |  | m³ |
| Total Building Volume Variance |  |  | 1000/2000 |  | m³ |
| Total Building Volume Standard Deviation |  |  | 1000/2000 |  | m³ |
| Green Spaces: Tree Cover and Normalised Difference Vegetation Index (NDVI) |  |  |  |  |  |
| Normalised Difference Vegetation Index (NDVI) | 14 | 300 m | None | Every 10 days (2013-2021) |  |
| Tree Cover | 13 | 30 m | 30/300/500/1000/3000/5000 | 2010 | % |
| Blue Spaces: Distance to Waterbodies |  |  |  |  |  |
| Surrounding Waterbodies | 15 | 30 m | 300/500/1000/3000/5000 | Yearly (1984-2021) |  |
| Surrounding Waterbodies Gaussian Filter |  |  |  |  | % |
| Light Exposure: Night-time Lights and Solar Radiation |  |  |  |  |  |
| Night-time Lights (NTL) | 16 | 1000 m | None | Yearly (1992-2018) |  |
| SARAH-2 | 17 | 0.05° | None | Monthly (2005-2020) | W/m2 |
| Solar Radiation | Julien Radoux | 300 m | None | Weekly (static) | W/m² |
| Elevation: Copernicus Digital Elevation Model (DEM) |  |  |  |  |  |
| Elevation | 18 | 30 m | 30/300/500/1000/3000/5000 | static with data from 2010-2015, 2019 | m |
| Elevation Range |  |  | 30/300/500/1000/3000/5000 |  | m |
| Elevation Standard Deviation |  |  | 30/300/500/1000/3000/5000 |  | m |
| Slope |  |  | 30/300/500/1000/3000/5000/7500 |  | ° |
| Aspect |  |  | None |  | ° |
| Terrain Ruggedness Index (TRI) |  |  | 90 |  |  |
| Roughness |  |  | 90 |  |  |

| Variable Name | Data Source | Spatial Resolution | Temporal Coverage | Units |
| --- | --- | --- | --- | --- |
| Cloud cover (total, high, medium and low clouds) | 19 | 0.25° | Hourly,<br>Monthly means and standard deviation |  |
| Temperature, 2 m |  |  | Hourly,<br>Diurnal range,<br>Daily and monthly mean, maximum and minimum | °C |
| Temperature, dewpoint |  |  | Hourly,<br>Daily and monthly diurnal range, mean, maximum and minimum<br>Monthly diurnal and mean standard deviation | °C |
| Wet-bulb globe temperature (WBGTmax) |  |  | Daily and monthly mean. Monthly standard deviation | °C |
| Precipitation volume |  |  | Hourly,<br>Daily and monthly mean. Monthly standard deviation | mm/d ay |
| Precipitation duration |  |  | Daily and monthly duration. Monthly standard deviation. Daily duration of maximum events [h] | h/day |
| Total number of wet days | | | Monthly total number of wet days [$\geq 1$mm] | days |
| Simple Precipitation Intensity Index | | | Mean precipitation on wet days [$\geq 1$mm] during the month | mm/d ay |
| Cloud cover (total, high, medium and low clouds) |  |  |  |  |
| TXx: Monthly maximum value of daily maximum temperature | 19 | 0.25° | Monthly | °C |
| TNx: Monthly maximum value of daily minimum temperature |  |  | Monthly | °C |
| TNn: Monthly minimum value of daily minimum temperature |  |  | Monthly | °C |
| TX90p: Percentage of warm days (>90<sup>th</sup> percentile) |  |  | Monthly | % |
| TN90p: Percentage of warm nights (>90<sup>th</sup> percentile) |  |  | Monthly | % |
| TX10p: Percentage of cold days (<10<sup>th</sup> percentile) |  |  | Monthly | % |
| TN10p: Percentage of cold nights (<10<sup>th</sup> percentile) |  |  | Monthly | % |
| Warm spell duration index |  |  | Yearly | days |
| Cold spell duration index |  |  | Yearly | days |
| WBG Tmax90p: Percentage of days with extreme WBG Tmax (>90<sup>th</sup> percentile) |  |  | Monthly | % |
| CWD: Maximum number of consecutive wet days |  |  | Monthly | days |
| Rx5D: Monthly highest 5-day precipitation amount |  |  | Monthly | mm |
| R10mm: Heavy precipitation days (>=10mm) |  |  | Monthly | days |
| R20mm: Very heavy precipitation days (>=20mm) |  |  | Monthly | days |
| R75pTOT: Moderate wet days per month (>=75<sup>th</sup> percentile) |  |  | Monthly | days |
| R95pTOT: Very wet days per month (>=95<sup>th</sup> percentile) |  |  | Monthly | days |
| Sea salt aerosol mixing ratios | 20 | 0.75° | 3-hourly, Monthly mean and standard deviation | kg/kg<sup>-1</sup> |
| Dust aerosol mixing ratios |  |  |  | kg/kg<sup>-1</sup> |
| Hydrophilic organic matter aerosol mixing ratio |  |  |  | kg/kg<sup>-1</sup> |
| Hydrophobic organic matter aerosol mixing ratio |  |  |  | kg/kg<sup>-1</sup> |
| Hydrophilic black carbon aerosol mixing ratio |  |  |  | kg/kg<sup>-1</sup> |
| Hydrophobic black carbon aerosol mixing ratio |  |  |  | kg/kg<sup>-1</sup> |
| Sulphate aerosol mixing ratio |  |  |  | kg/kg<sup>-1</sup> |
| Nitrogen dioxide mass mixing ratio (NO<sub>2</sub>) |  |  |  | kg/kg<sup>-1</sup> |
| Sulphur dioxide mass mixing ratio (SO<sub>2</sub>) |  |  |  | kg/kg<sup>-1</sup> |
| Total column carbon monoxide (CO) |  |  |  | kg/m<sup>-2</sup> |
| Total column formaldehyde (CH<sub>2</sub>O) |  |  |  | kg/m<sup>-2</sup> |
| Total column methane (CH<sub>4</sub>) |  |  |  | kg/m<sup>-2</sup> |
| Total column ozone (O<sub>3</sub>) |  |  |  | kg/m<sup>-2</sup> |
| Black carbon aerosol optical depth (AOD) at 550 nm |  |  |  |  |
| Dust AOD at 550 nm |  |  |  |  |
| Organic matter AOD at 550 nm |  |  |  |  |
| Sea salt AOD at 550 nm |  |  |  |  |
| Sulphate AOD at 550 nm |  |  |  |  |
| Total AOD at 550 nm |  |  |  |  |
| Particulate matter d < 1 µm (PM<sub>1</sub>) |  |  |  | kg/m<sup>-3</sup> |
| Particulate matter d < 2.5 µm (PM<sub>2.5</sub>) |  |  |  | kg/m<sup>-3</sup> |
| Particulate matter d < 10 µm (PM<sub>10</sub>) |  |  |  | kg/m<sup>-3</sup> |

## Supplement B: Exploratory exposure–depression gradients across environmental indicators

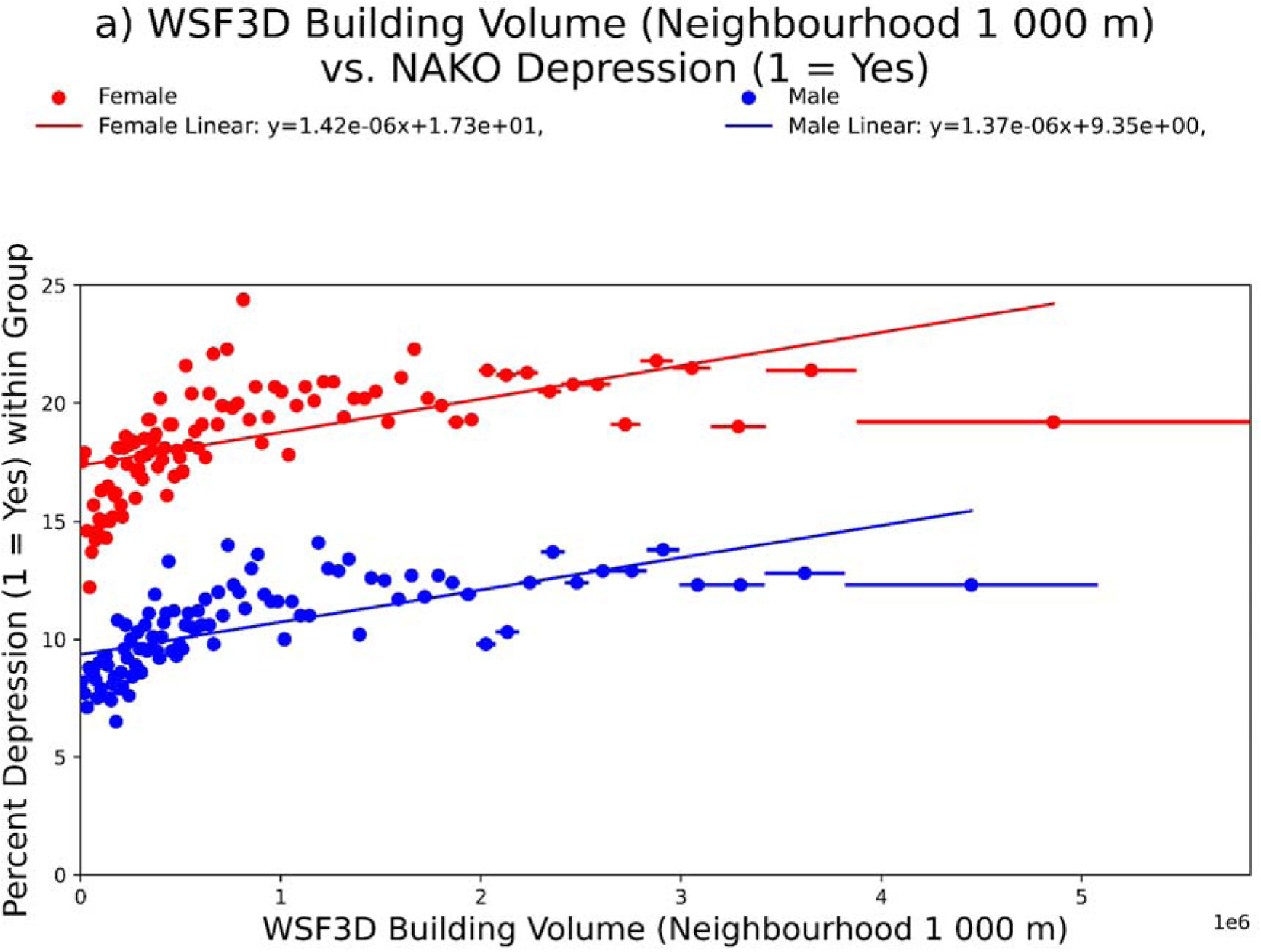
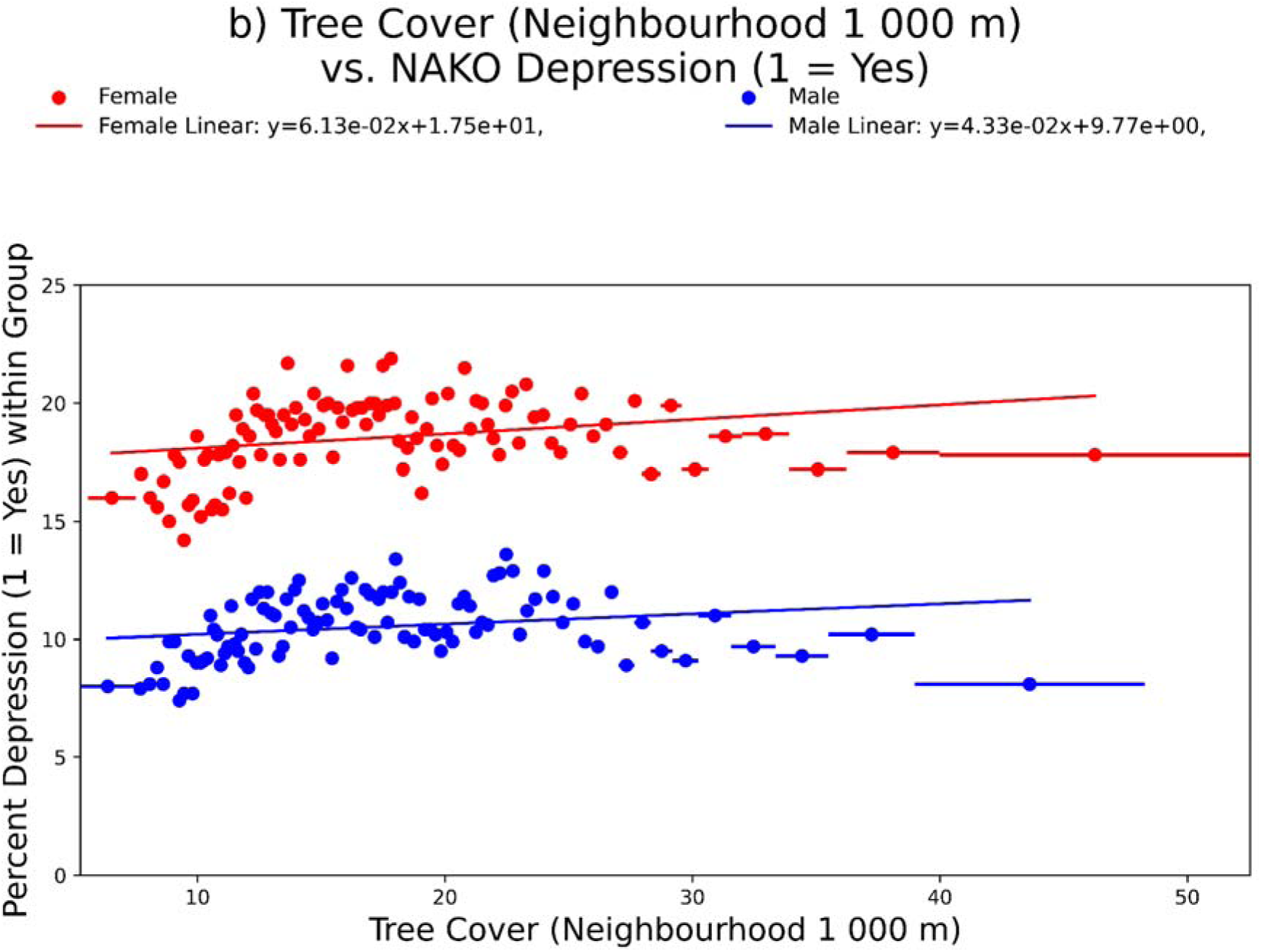
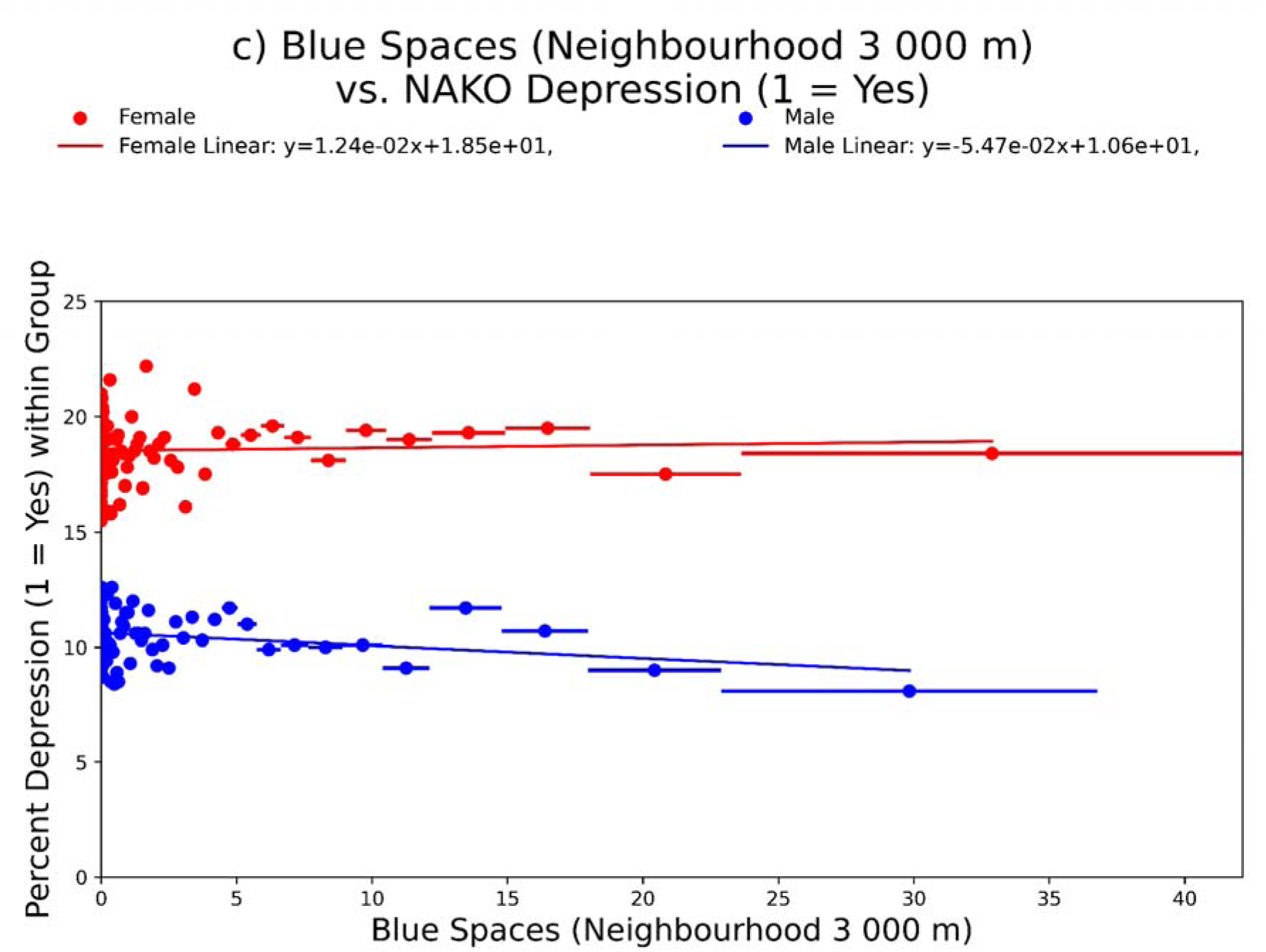
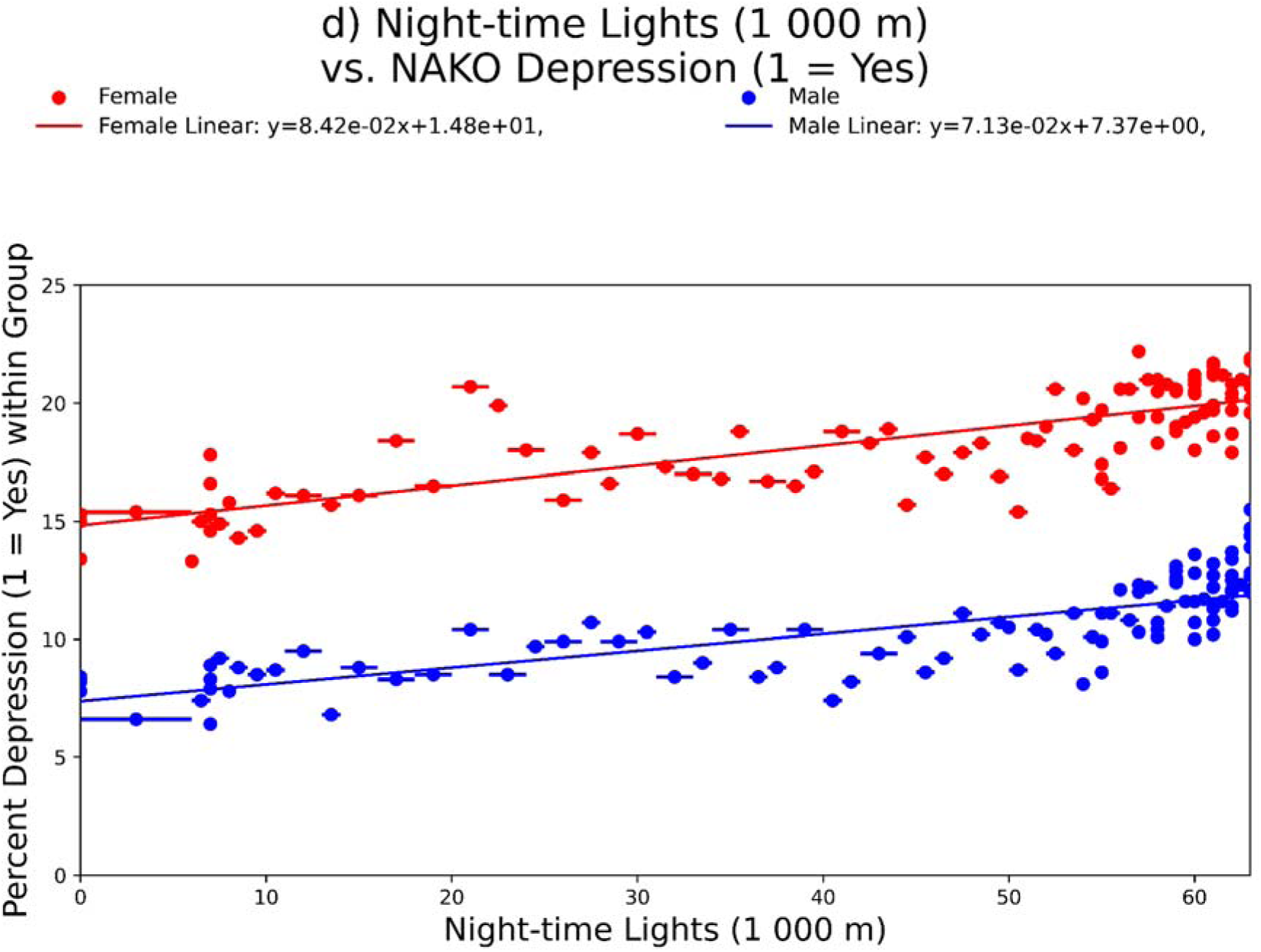
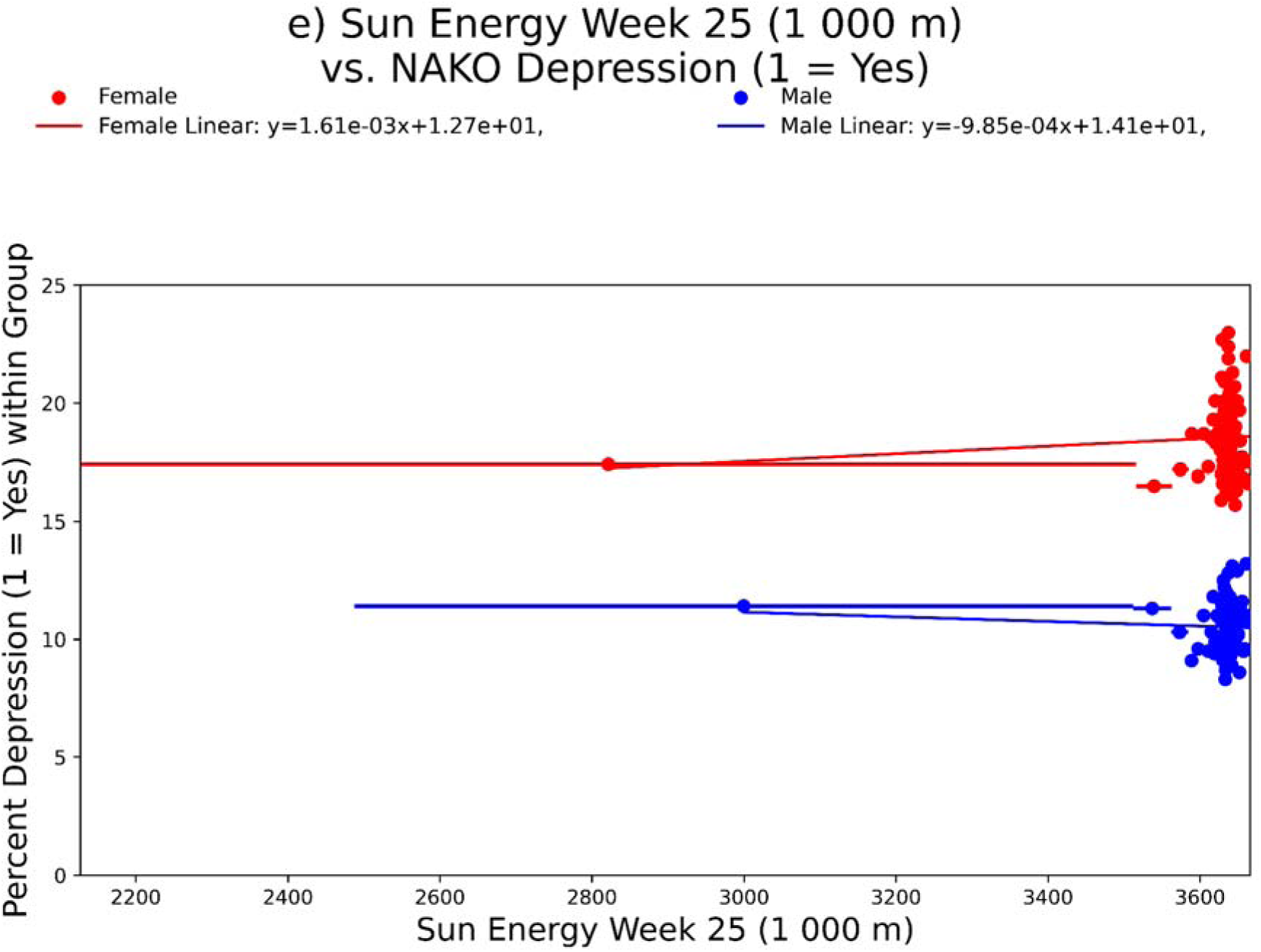
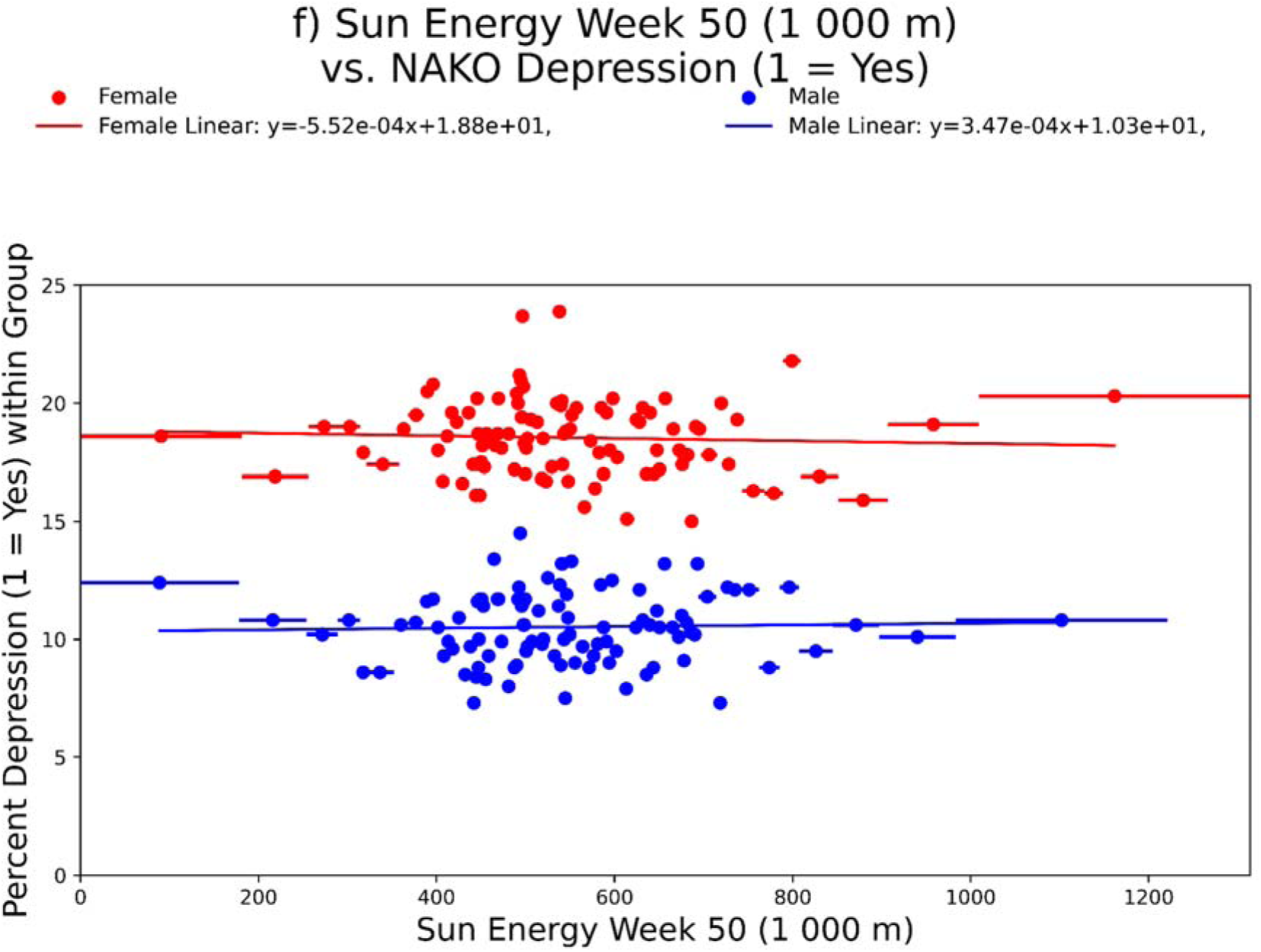
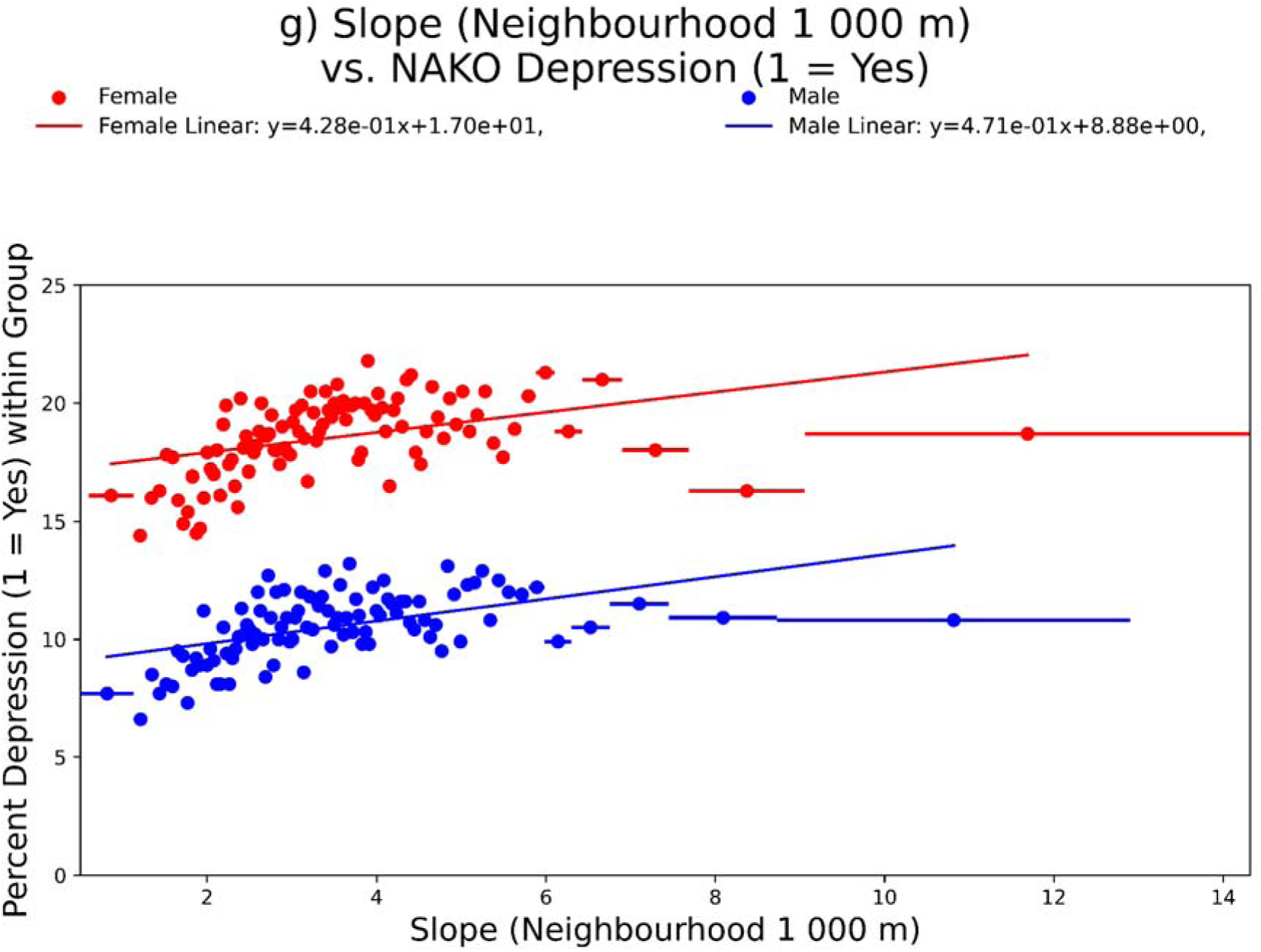
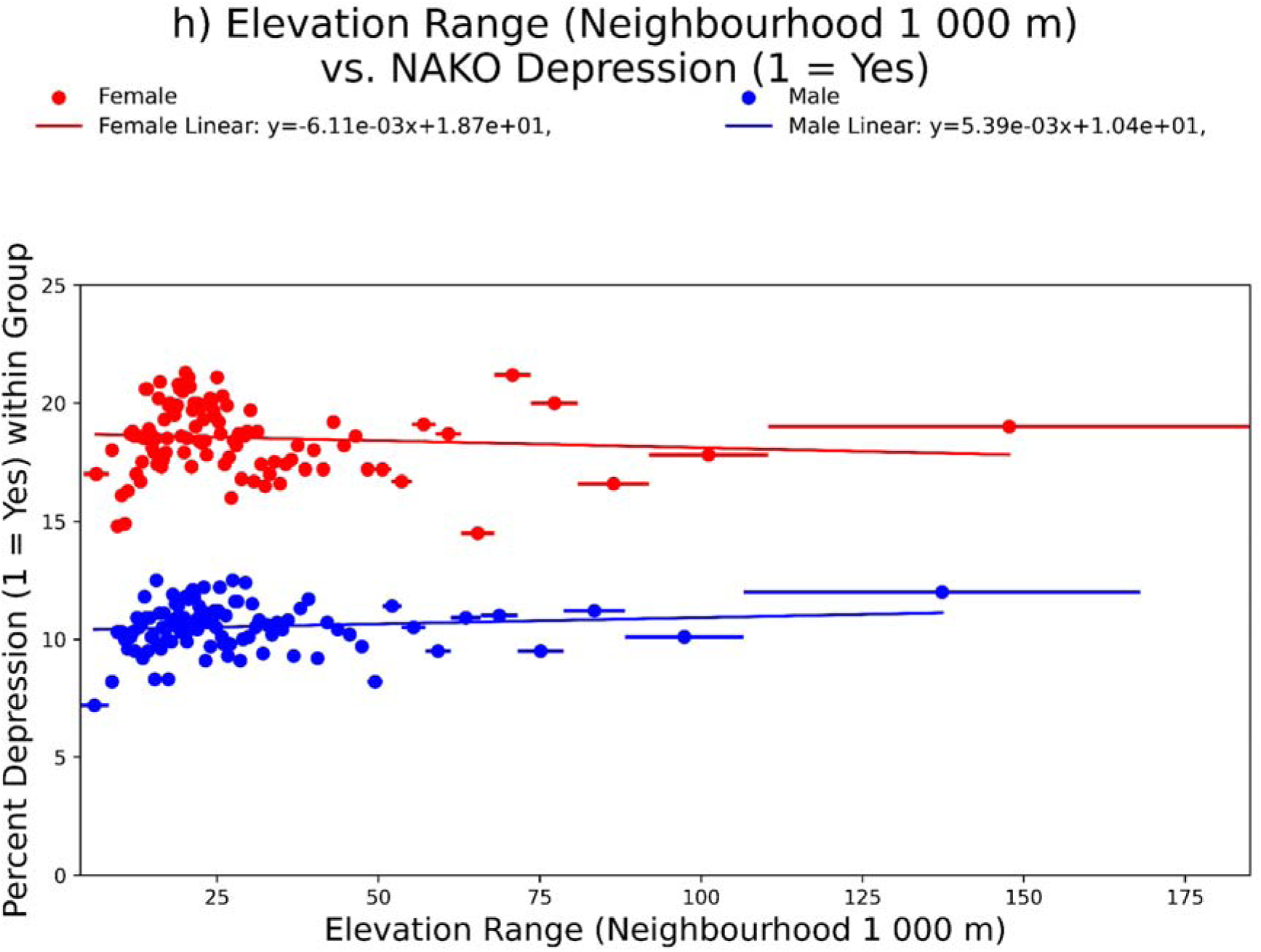

## References

1. Polemiti, E., Hese, S., Schepanski, K., Yuan, J. & Schumann, G. How does the macroenvironment influence brain and behaviour-a review of current status and future perspectives. Molecular psychiatry; 10.1038/s41380-024-02557-x (2024).

2. Reuben, A. et al. The Interplay of Environmental Exposures and Mental Health: Setting an Agenda. Environmental health perspectives 130, 25001; 10.1289/EHP9889 (2022).

3. Nobile, F., Forastiere, A., Michelozzi, P., Forastiere, F. & Stafoggia, M. Long-term exposure to air pollution and incidence of mental disorders. A large longitudinal cohort study of adults within an urban area. Environment International 181, 108302; 10.1016/j.envint.2023.108302 (2023).

4. Qiu, X. et al. Associations of short-term exposure to air pollution and increased ambient temperature with psychiatric hospital admissions in older adults in the USA: a case-crossover study. The Lancet. Planetary health 6, e331–e341; 10.1016/S2542-5196(22)00017-1 (2022).

5. Engemann, K. et al. Residential green space in childhood is associated with lower risk of psychiatric disorders from adolescence into adulthood. Proceedings of the National Academy of Sciences of the United States of America 116, 5188–5193; 10.1073/pnas.1807504116 (2019).

6. Keijzer, C. de, Bauwelinck, M. & Dadvand, P. Long-Term Exposure to Residential Greenspace and Healthy Ageing: a Systematic Review. Current environmental health reports 7, 65–88; 10.1007/s40572-020-00264-7 (2020).

7. Lederbogen, F. et al. City living and urban upbringing affect neural social stress processing in humans. Nature 474, 498–501; 10.1038/nature10190 (2011).

8. Obradovich, N., Migliorini, R., Paulus, M. P. & Rahwan, I. Empirical evidence of mental health risks posed by climate change. Proceedings of the National Academy of Sciences of the United States of America 115, 10953–10958; 10.1073/pnas.1801528115 (2018).

9. Vanoli, J. et al. Correction: Reconstructing individual-level exposures in cohort analyses of environmental risks: an example with the UK Biobank. Journal of exposure science & environmental epidemiology 35, 692; 10.1038/s41370-025-00771-5 (2025).

10. German National Cohort Consortium. The German National Cohort: aims, study design and organization. European journal of epidemiology 29, 371–382; 10.1007/s10654-014-9890-7 (2014).

11. Peters, A. et al. Framework and baseline examination of the German National Cohort (NAKO). European journal of epidemiology 37, 1107–1124; 10.1007/s10654-022-00890-5 (2022).

12. Esch, T. et al. World Settlement Footprint 3D - A first three-dimensional survey of the global building stock. Remote Sensing of Environment 270, 112877; 10.1016/j.rse.2021.112877 (2022).

13. Hansen, M. C. et al. High-resolution global maps of 21st-century forest cover change. *Science (New York*, N.Y.) 342, 850–853; 10.1126/science.1244693 (2013).

14. Sterckx, S. et al. The PROBA-V mission: image processing and calibration. International Journal of Remote Sensing 35, 2565–2588; 10.1080/01431161.2014.883094&sa=D&source=docs&ust=1715602796580134&usg=AOvVaw1abKuVnr4pQTOPHG90A7aD (2014).

15. Pekel, J.-F., Cottam, A., Gorelick, N. & Belward, A. S. High-resolution mapping of global surface water and its long-term changes. Nature 540, 418–422; 10.1038/nature20584 (2016).

16. Li, X., Zhou, Y., Zhao, M. & Zhao, X. A harmonized global nighttime light dataset 1992-2018. Scientific data 7, 168; 10.1038/s41597-020-0510-y (2020).

17. Muellejans, H. et al. State-of-the-art assessment of solar energy technologies. 1831-9424; 10.2760/735053 (2022).

18. European Union, ESA, DLR e.V., Airbus Defence and Space GmbH. Copernicus DEM (2022).

19. Hersbach, H. et al. The ERA5 global reanalysis. Quart J Royal Meteoro Soc 146, 1999–2049; 10.1002/qj.3803 (2020).

20. Inness, A. et al. The CAMS reanalysis of atmospheric composition. Atmos. Chem. Phys. 19, 3515–3556; 10.5194/acp-19-3515-2019 (2019).

21. The European Space Agency through its establishment the European Space Research Institute (ESRIN). Copernicus Data Space Ecosystem Documentation Portal. Available at https://documentation.dataspace.copernicus.eu/.

22. Mauz, E. et al. Cohort profile: KiGGS cohort longitudinal study on the health of children, adolescents and young adults in Germany. International journal of epidemiology 49, 375–375k; 10.1093/ije/dyz231 (2020).

23. Magnus, P. et al. Cohort Profile Update: The Norwegian Mother and Child Cohort Study (MoBa). International journal of epidemiology 45, 382–388; 10.1093/ije/dyw029 (2016).

24. Sudlow, C. et al. UK biobank: an open access resource for identifying the causes of a wide range of complex diseases of middle and old age. PLoS medicine 12, e1001779; 10.1371/journal.pmed.1001779 (2015).

25. Schumann, G. et al. The IMAGEN study: reinforcement-related behaviour in normal brain function and psychopathology. Molecular psychiatry 15, 1128–1139; 10.1038/mp.2010.4 (2010).

26. Xu, Q. et al. CHIMGEN: a Chinese imaging genetics cohort to enhance cross-ethnic and cross-geographic brain research. Molecular psychiatry 25, 517–529; 10.1038/s41380-019-0627-6 (2020).

27. Qu, D. et al. Balancing the 24-hour lifestyle: A population-based study on reducing mental health problems, self-injurious thoughts and behaviors among adolescents. Journal of affective disorders 387, 119451; 10.1016/j.jad.2025.119451 (2025).

28. Sharma, E. et al. Consortium on Vulnerability to Externalizing Disorders and Addictions (cVEDA): A developmental cohort study protocol. BMC psychiatry 20, 2; 10.1186/s12888-019-2373-3 (2020).

29. Li, S. et al. Sex difference in incidence of major depressive disorder: an analysis from the Global Burden of Disease Study 2019. Annals of general psychiatry 22, 53; 10.1186/s12991-023-00486-7 (2023).

30. Chen, T.-H. K. et al. Higher depression risks in medium- than in high-density urban form across Denmark. Science advances 9, eadf3760; 10.1126/sciadv.adf3760 (2023).

31. Xu, J. et al. Effects of urban living environments on mental health in adults. Nature medicine 29, 1456–1467; 10.1038/s41591-023-02365-w (2023).

32. Liu, Z. et al. Green space exposure on depression and anxiety outcomes: A meta-analysis. Environmental research 231, 116303; 10.1016/j.envres.2023.116303 (2023).

33. Geary, R. S. et al. Ambient greenness, access to local green spaces, and subsequent mental health: a 10-year longitudinal dynamic panel study of 2·3 million adults in Wales. The Lancet. Planetary health 7, e809–e818; 10.1016/S2542-5196(23)00212-7 (2023).

34. White, M. P. et al. Associations between green/blue spaces and mental health across 18 countries. Scientific reports 11, 8903; 10.1038/s41598-021-87675-0 (2021).

35. Wright, K. et al. A qualitative exploration of the contribution of blue space to welllJbeing in the lives of people with severe mental illness. People and Nature 6, 849–864; 10.1002/pan3.10620 (2024).

36. Liu, Y. et al. Longitudinal associations between neighbourhood physical environments and depressive symptoms of older adults in Hong Kong: The moderating effects of terrain slope and declining functional abilities. Health & place 70, 102585; 10.1016/j.healthplace.2021.102585 (2021).

37. Evans, G. W. The built environment and mental health. Journal of urban health : bulletin of the New York Academy of Medicine 80, 536–555; 10.1093/jurban/jtg063 (2003).

38. Zijlema, W. et al. Cities and mental health: The role of the built environment, and environmental and lifestyle factors in Barcelona. Environmental pollution (Barking, Essex : 1987) 346, 123559; 10.1016/j.envpol.2024.123559 (2024).

39. Barton, J. & Rogerson, M. The importance of greenspace for mental health. BJPsych international 14, 79–81; 10.1192/s2056474000002051 (2017).

40. Triguero-Mas, M. et al. Natural outdoor environments and mental and physical health: Relationships and mechanisms. Environment International 77, 35–41; 10.1016/j.envint.2015.01.012 (2015).

41. McDougall, C. W. et al. Neighbourhood blue space and mental health: A nationwide ecological study of antidepressant medication prescribed to older adults. Landscape and Urban Planning 214, 104132; 10.1016/j.landurbplan.2021.104132 (2021).

42. Liu, B.-P. et al. Exposure to residential green and blue space and the natural environment is associated with a lower incidence of psychiatric disorders in middle-aged and older adults: findings from the UK Biobank. BMC medicine 22, 15; 10.1186/s12916-023-03239-1 (2024).

43. Pearson, A. L. et al. Effects of freshwater blue spaces may be beneficial for mental health: A first, ecological study in the North American Great Lakes region. PloS one 14, e0221977; 10.1371/journal.pone.0221977 (2019).

44. Völker, S. & Kistemann, T. Developing the urban blue: Comparative health responses to blue and green urban open spaces in Germany. Health & place 35, 196–205; 10.1016/j.healthplace.2014.10.015 (2015).

45. Lehmler, S. et al. Closing the loop between environment, brain and mental health: how far we might go in real-life assessments? Current opinion in psychiatry 37, 301–308; 10.1097/YCO.0000000000000941 (2024).

46. Vohland, K. et al. The Science of Citizen Science (Springer International Publishing, Cham, 2021).

47. Dormann, C. F. et al. Methods to account for spatial autocorrelation in the analysis of species distributional data: a review. Ecography 30, 609–628; 10.1111/j.2007.0906-7590.05171.x (2007).

48. Smith, M. J., Phillips, R. V., Luque-Fernandez, M. A. & Maringe, C. Application of targeted maximum likelihood estimation in public health and epidemiological studies: a systematic review. Annals of epidemiology 86, 34–48.e28; 10.1016/j.annepidem.2023.06.004 (2023).

49. Keil, A. P. et al. A Quantile-Based g-Computation Approach to Addressing the Effects of Exposure Mixtures. Environmental health perspectives 128, 47004; 10.1289/EHP5838 (2020).

